# Identifying the potential role of insomnia on multimorbidity: A Mendelian randomization phenome-wide association study in UK Biobank

**DOI:** 10.1101/2022.01.11.22269005

**Authors:** Mark J Gibson, Deborah A Lawlor, Louise AC Millard

**Author notes:** these authors contributed equally (Deborah A Lawlor, Louise AC Millard).

## Abstract

**Objectives:** To identify the breadth of potential causal effects of insomnia on health outcomes and hence its possible role in multimorbidity.

**Design:** Mendelian randomisation (MR) Phenome-wide association study (MR-PheWAS) with two-sample Mendelian randomisation follow-up.

**Setting:** Individual data from UK Biobank and summary data from a number of genome-wide association studies.

**Participants:** 336,975 unrelated white-British UK Biobank participants.

**Exposures:** Standardised genetic risk of insomnia for the MR-PheWAS and genetically predicted insomnia for the two-sample MR follow-up, with insomnia instrumented by a genetic risk score (GRS) created from 129 single-nucleotide polymorphisms (SNPs).

**Main outcomes measures:** 11,409 outcomes from UK Biobank extracted and processed by an automated pipeline (PHESANT). Potential causal effects (i.e., those passing a Bonferroni-corrected significance threshold) were followed up with two-sample MR in MR-Base, where possible.

**Results:** 437 potential causal effects of insomnia were observed for a number of traits, including anxiety, stress, depression, mania, addiction, pain, body composition, immune, respiratory, endocrine, dental, musculoskeletal, cardiovascular and reproductive traits, as well as socioeconomic and behavioural traits. We were able to undertake two-sample MR for 71 of these 437 and found evidence of causal effects (with directionally concordant effect estimates across all analyses) for 25 of these. These included, for example, risk of anxiety disorders (OR=1.55 [95% confidence interval (CI): 1.30, 1.86] per category increase in insomnia), diseases of the oesophagus/stomach/duodenum (OR=1.32 [95% CI: 1.14, 1.53]) and spondylosis (OR=1.57 [95% CI: 1.22, 2.01]).

**Conclusion:** Insomnia potentially causes a wide range of adverse health outcomes and behaviours. This has implications for developing interventions to prevent and treat a number of diseases in order to reduce multimorbidity and associated polypharmacy.

What this paper adds

What is already known on this topic

- Insomnia symptoms are widespread in the population and insomnia is the second most common mental health disorder after anxiety disorders.
- Insomnia might have broad and numerous effects on health and multimorbidity, but both observational and Mendelian randomisation studies have focused on select hypothesised associations/effects rather than taking a systematic hypothesis-free approach across many health outcomes.

What this study adds

- This study uses a hypothesis-free approach to systematically identify causal effects of insomnia on 11,409 health outcomes; it identified 437 potential causal effects and for 71 that could be followed-up with two-sample MR, 25 showed evidence of a causal effect and were directionally consistent across all analyses.
- These findings identify potential routes for developing interventions to prevent and treat a number of diseases and reduce multimorbidity and polypharmacy.

## Introduction

While there is still much debate over the exact purpose of sleep, it is clear that sleep is vital for healthy functioning and likely to be multifaceted. Experiments on rats have suggested that sleep is linked to antioxidative enzyme levels in the brain which regulate the levels of reactive oxygen species (by-products of the metabolization of oxygen which damage cells) (1). It has also been proposed that sleep is vital for the consolidation of information, learning and memory (2, 3). Insomnia is defined as regular dissatisfaction with the quality or quantity of sleep for a prolonged period and includes difficulty initiating or maintaining sleep (4). Evidence suggests that 6-7% of the European population have a diagnosis of insomnia, while 33-37% self-report having insomnia symptoms (5-7). It is the second most prevalent mental health disorder (after anxiety disorder) and is more common in women and the elderly (6, 7). Multimorbidity, defined as patients living with two or more chronic health conditions, is associated with polypharmacy, poor quality of life and premature mortality (8, 9). It is increasingly recognised as a threat to global health and identifying potential causes of multimorbidity is a research priority (10). Given the high prevalence of insomnia symptoms, and their association with many diseases (including depression (11), diabetes (12), hypertension (13), dementia (14) and cardiovascular disease (15, 16)), insomnia could be a cause of multimorbidity.

However, associations with disease outcomes may not be due to a causal effect of insomnia on the outcome and could reflect residual confounding or reverse causality from undiagnosed prevalent disease (17). Furthermore, studies to date have focused on hypothesised selected outcomes, predominantly mental, neurocognitive and cardiometabolic outcomes, rather than systematically, using a hypothesis free approach, searching for potential causal effects across a wide range of health and disease outcomes.

Mendelian randomisation (MR) is a method used for testing causal relationships, that generally uses genetic variants that are robustly associated with the exposure of interest as instrumental variables (IV) (18). MR is typically less prone to confounding of the exposure outcome association and reverse causation than conventional observational epidemiology; as genetic variation is determined at conception it cannot be altered by disease status (19). However, it has other potential sources of bias, in particular those due to weak instruments, confounding of the instrument-outcome association and horizontal pleiotropy (20) (the core assumptions of MR have been previously reported in detail (21)). Previous MR studies have provided some evidence that insomnia may lead to heavier substance use (22, 23), increased BMI, an increased risk of type 2 diabetes (24), cardiovascular diseases (25), autism spectrum disorder, bipolar disorder (26), pain (27), and major depressive disorder (28), and also increased levels of an inflammatory marker (glycoprotein acetyls) and citrate, but with no strong evidence that it causes widespread metabolic disruption across 115 metabolic markers (29). However, insomnia may have a causal effect on many health outcomes beyond those already studied. If insomnia is a cause of multimorbidity then insomnia treatments, such as the UK national institute for health and care excellence (NICE) guideline (30) recommended cognitive behavioural therapy-Insomnia (31), would prevent a range of other adverse health outcomes.

MR-PheWAS combines both MR and Phenome Wide Association Studies (PheWAS) (32) to explore causal relationships with many phenotypes in a hypothesis-free manner (33). To our knowledge only one previous study has undertaken an MR-PheWAS of insomnia (34). In that study the automated tool PhenoScanner (35) was used to explore causal effects of insomnia on 179 outcomes (34). It identified 478 potential causal effects (using on a p-value threshold of 5 × 10^−8^) including on adiposity, mental health, musculoskeletal, respiratory/allergic and reproductive phenotypes. However, that MR-PheWAS was part of an illustrative example in a methodological paper fousing on addressing one of the MR assumptions, and none of the potential causal effects were explored further with replication or sensitivity analyses. Here we use an open-source software package called PHESANT (36) to conduct a large-scale MR-PheWAS in UK Biobank (37, 38), to search for novel causal effects of insomnia on health outcomes. The authors followed the STROBE-MR reporting guidelines when writing this paper (39) and this study was not pre-registered.

## Methods

### Study Population

We used data from UK Biobank, a large prospective cohort study (dataset ID 43017 of UK Biobank application 16729, phenotypic data extracted on 24/02/2021). UK Biobank recruited 503,325 adults aged between 37–73 years. They were recruited between 2006 and 2007 and attended one of 22 test centres across the UK. Of the 503,325 participants, genetic data (40) was successfully obtained for 487,406 participants (Supplementary Text). Participants were then excluded from this sample if they did not meet the genetic quality control (41), they were not of white-British ancestry, they were not part of the maximal subset of individuals not related to any other individual to the third degree or higher, or they had since withdrawn their consent (as of 09/08/2021). The remaining 336,975 participants were included in the MR-PheWAS (See Figure 1 for a flow diagram).

**Figure 1.**
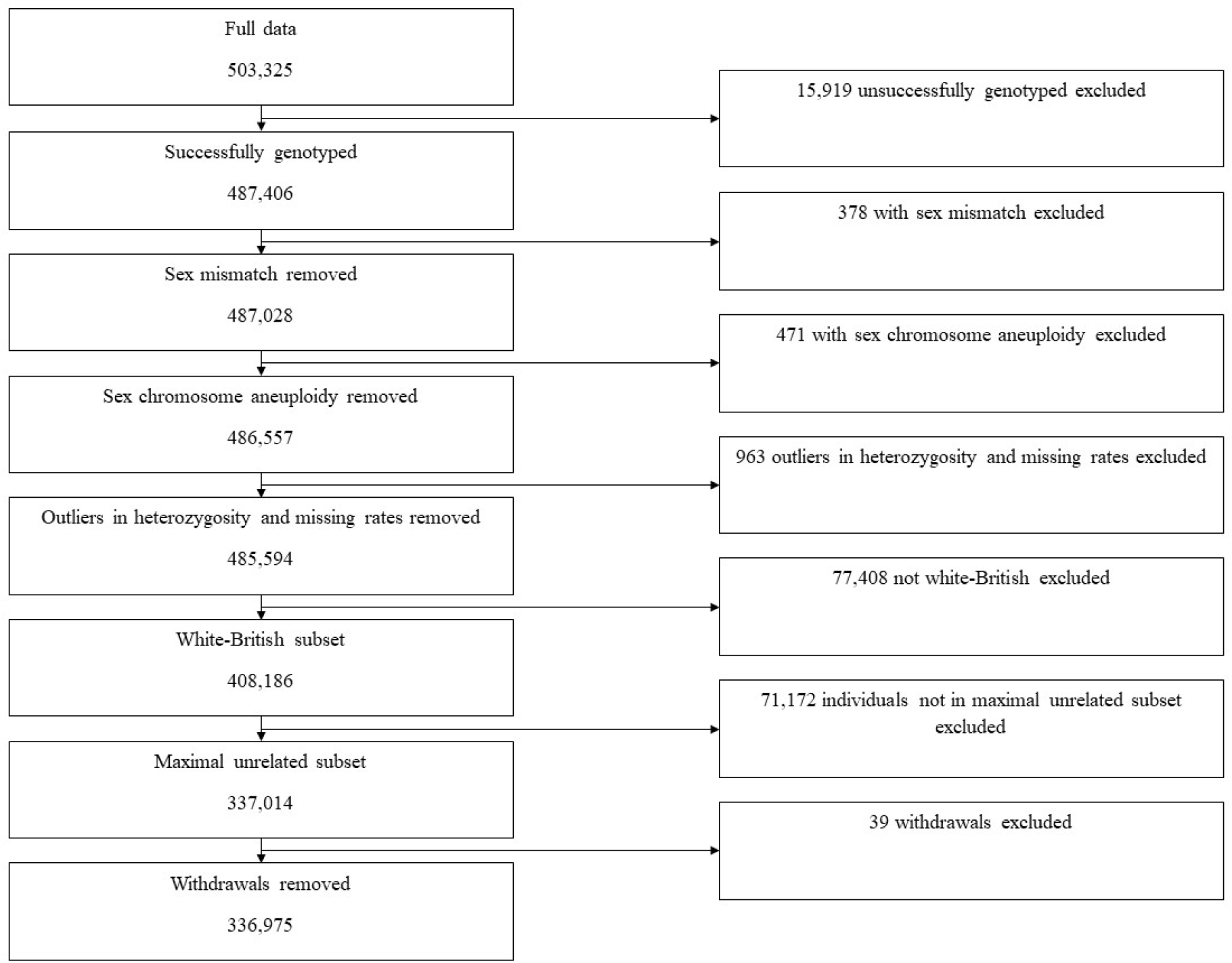
Flow chart of participant inclusion.

### Genetic Risk Score

We generated a weighted genetic risk score (GRS) using 129 independent single-nucleotide polymorphisms (SNPs) previously identified to associate with insomnia (Supplementary text) at GWAS significance (with p< 5 × 10^−8^) in 23andMe, Inc. (24). These data were requested from 23andMe as they were not provided in the original GWAS paper. SNPs were weighted by their per-allele association with insomnia in the original GWAS (Supplementary Table S1). We used a linkage disequilibrium (LD) threshold of R^2^>0.001 to clump the GWAS significant SNPs into independent SNPs. LD was calculated in the 1000 Genomes European data (42) and the TwoSampleMR (MR-base) R package v0.5.6 (43) was used to clump GWAS significant SNPs into independent SNPs. One SNP (rs28458909) was not available in UK Biobank and thus was replaced by a proxy (rs28780988) that was in close LD (R^2^ = 1). All palindromic SNPs had an effect allele frequency (EAF) falling below 0.49 or above 0.51 in UK Biobank and 23andMe and therefore could be harmonised.

### Outcomes

11,409 outcome variables were derived and analysed using PHESANT (36). Outcomes included those obtained from responses to baseline and follow-up questionnaires, baseline assessments such as weight, height, blood pressure and DXA scan bone density measurements, follow-up assessments such as accelerometer measurements and a range of different scans (including brain and cardiac scans), biomarker measures from blood or urine samples and outcomes from linkage to primary and secondary care, and the national cancer and death registers. In order to summarise our overall findings from the MR-PheWAS, outcomes were assigned to categories (e.g., Mental Health) based on their UK Biobank category (e.g., Online follow-up > Mental health > Anxiety). Measurements that were not health related outcomes were assigned to the Auxiliary Variables category. These included outcomes such as hospital administration records and procedural metrics such as the length of time taken to complete the touch screen questionnaire. Individual sleep variables from the mental health and physical health categories were then reassigned to a sleep category and medication variables in the physical health category that were for mental disorders were reassigned to the mental health category. We then manually assigned outcomes in these two categories to subcategories. Supplementary Table S3 shows which category and subcategory each UK Biobank category is assigned to.

### PheWAS Analysis

The PHESANT v1.0 package was used for the MR-PheWAS. We adjusted for age at assessment, sex and the top 10 genetic principal components to control for populations stratification (44). A complete case analysis was undertaken by PHESANT meaning participant numbers differ between outcomes and we chose to exclude outcomes with less than 100 cases. PHESANT derives outcomes from the UK Biobank data and defines whether they are continuous, binary, ordered categorical or unordered categorical and tests the association with a trait of interest, in our case the insomnia GRS, using linear (using inverse normal rank transformed data to ensure a normal distribution), logistic, ordered logistic, and multinomial logistic regression, respectively. The results are presented as difference in mean SD of inverse rank normal transformed continuous outcomes and odds ratio (OR) for categorical outcomes, per 1 standard deviation (SD) increase in the weighted GRS. We defined *potential causal effects* as any insomnia GRS-outcome association that passed the Bonferroni-corrected significance threshold of 4.38×10^−6^ (0.05/11,409) in the MR-PheWAS. The less conservative false discovery rate (FDR) correction was also calculated and reported but was not used to identify potential causal effects for follow-up.

### Sensitivity Analysis

As the SNPs used to construct the GRS in the main analysis are not replicated there is a higher chance that spurious SNPs could have been falsely detected, increasing the chance that the GRS may contain horizontally pleiotropic SNPs. We created two sensitivity analysis GRS which used SNPs which were replicated in UK Biobank, meaning the presence of spurious SNPs is less likely. However, as the MR-PheWAS was conducted in UK Biobank this overlap between the selection and test sample could introduce bias through overfitting or winner’s curse.

These GRSs included 111 SNPs (Supplementary Table S4) identified in a meta-analysis GWAS of both 23andMe and UK Biobank (24). The first sensitivity GRS (S1) weighted SNPs by the per-allele association with insomnia from the pooled UK Biobank and 23andMe analyses. The second sensitivity GRS (S2) weighted by the 23andMe per allele associations with insomnia. While 114 independent SNPs were identified in the original GWAS 3 SNPs were removed for the following reasons. One SNP (rs9540729) was palindromic and had an EAF in 23andMe between 0.49 and 0.51 meaning it could not be aligned with the UK Biobank data. Therefore, it was excluded from both scores for consistency. Two SNPs, rs77641763 and rs117630493, had point estimates in the meta-analysis GWAS which were in the opposite direction to the 23andMe GWAS, and so were also removed. Of the SNPs included in the score, 38 were also used in the main GRS. The authors of the original GWAS used UK Biobank data to calculate LD and an LD threshold of R^2^>0.001 was used to define independent SNPs.

### Follow-up two-sample MR

We undertook follow-up analyses using two-sample MR for all outcomes for which the association with the GRS was identified as a potential causal effect of insomnia. The purpose of this was to confirm the reliability of the potential causal effects identified in the MR-PheWAS and to provide a causal estimate. The TwoSampleMR (MR-base) v0.5.6 (43) was used to conduct the follow-up. It was decided *a priori* that outcomes included in the auxiliary variables or sleep categories would not be followed-up. First, we conducted an automated search for relevant GWAS using pre-specified search terms for each outcome. The search automatically excluded GWAS that included solely UK Biobank data, included non-European populations or stratified by sex, based on the meta-data included in the MR-Base database. Of the remaining GWAS, we excluded those that did not match a follow-up outcome on manual inspection, those for which the origins of the data used could not be determined, and those that used UK Biobank or 23andMe data. If the only GWAS available for a particular outcome included UK Biobank or 23andMe data (but did not only include UK Biobank or 23andMe data) we undertook follow-up in those GWAS and report the extent of overlap between the two samples. Of the remaining GWAS we then chose the most suitable for a given trait, that was either of the most suitable match in terms of the trait used or where multiple GWAS had suitable traits we chose the one with the larger sample size.

The two-sample MR analysis used the 129 SNPs used to construct the GRS in the main analysis. The SNP-insomnia associations were extracted from the 23andMe/UK Biobank meta-analysis GWAS summary data (24), and the SNP-outcome associations were extracted from the GWAS for each outcome. The TwoSampleMR (MR-base) package attempted to harmonise SNPs and excluded those it could not (e.g. if a suitable proxy cannot be found for missing SNPs or if SNPs were palindromic with allele frequencies near to 0.5). We used the inverse-variance weighted (IVW) method for our main two-sample MR analyses (45), and weighted median regression MR (46) and MR-Egger (47) as sensitivity analyses to explore potential bias due to unbalanced horizontal pleiotropy. Weighted median MR is unbiased when less than 50% of the weight is made of horizontally pleiotropic SNPs, while MR-Egger is unbiased when the magnitude of horizontal pleiotropy of the included SNPs is not proportional to the effect of each SNP on the exposure. All code can be found at https://github.com/MRCIEU/PHESANT-MR-PheWAS-Insomniav1.0.

## Results

### MR-PheWAS

The insomnia GRS was associated with an increased risk of insomnia in UK Biobank: Odds Ratio (OR) of report of usually versus never/rarely/sometimes having trouble falling or staying asleep = 1.08 [95% Confidence Interval (CI): 1.07, 1.09] per one standard deviation higher GRS (*p*=3.59×10^−84^, McFadden’s pseudo R^2^=0.01). See Supplementary Figure S1 for the association of each SNP with insomnia.

Of the 11,409 associations included in the MR-PheWAS, 437 were identified as potential causal effects (Supplementary Table S3). These included anxiety, stress, depression, mania, addiction, pain, body composition, immune, respiratory, endocrine, dental, musculoskeletal, cardiovascular and reproductive traits, as well as socioeconomic and behavioural traits. Figure 2 shows the proportion of potential causal effects of insomnia by broad categories of outcomes. For associations between insomnia and mental health outcomes 96 of 301 (32%) were identified as potential causal effects. There were higher proportions of these in 10 out of 17 of the mental health subcategories (Figure 3), including depression (38%), anxiety (48%), general (33%), well-being (87%), suicide and self-harm (24%), and mania (19%). Examples of adverse potential causal effects on mental health outcomes are having seen a doctor for nerves, anxiety, tension or depression (OR=1.06 per 1SD higher weighted GRS [95% CI: 1.05, 1.07]), neuroticism (OR=1.05, [95% CI: 1.04, 1.05]), mood (OR=1.05, [95% CI: 1.04, 1.05]) and the frequency of depressed moods in the last two weeks (OR=1.05, [95% CI: 1.04, 1.06]). By contrast none of the outcomes in subcategories of psychosis, personality disorders, organic brain disorders, eating disorders, developmental disorders or behavioural syndromes had associations with the GRS which were identified as potential causal effects.

**Figure 2.**
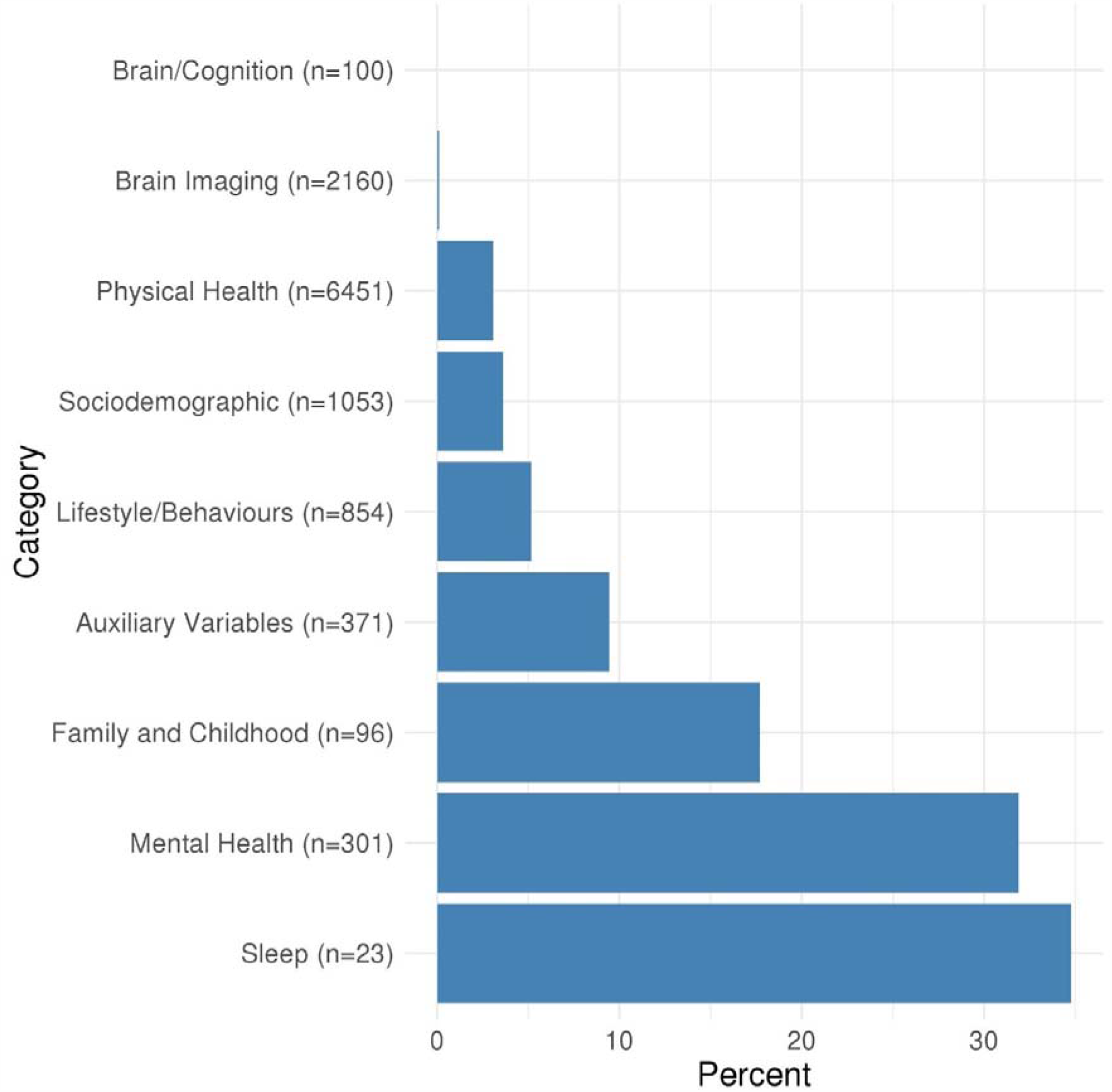
Proportion of potential causal effects of insomnia on outcomes within different categories. n is the total number of outcomes in the category. Supplementary Table S3 gives the category for each outcome. Results shown in this figure are also provided in Supplementary Table S4.

**Figure 3.**
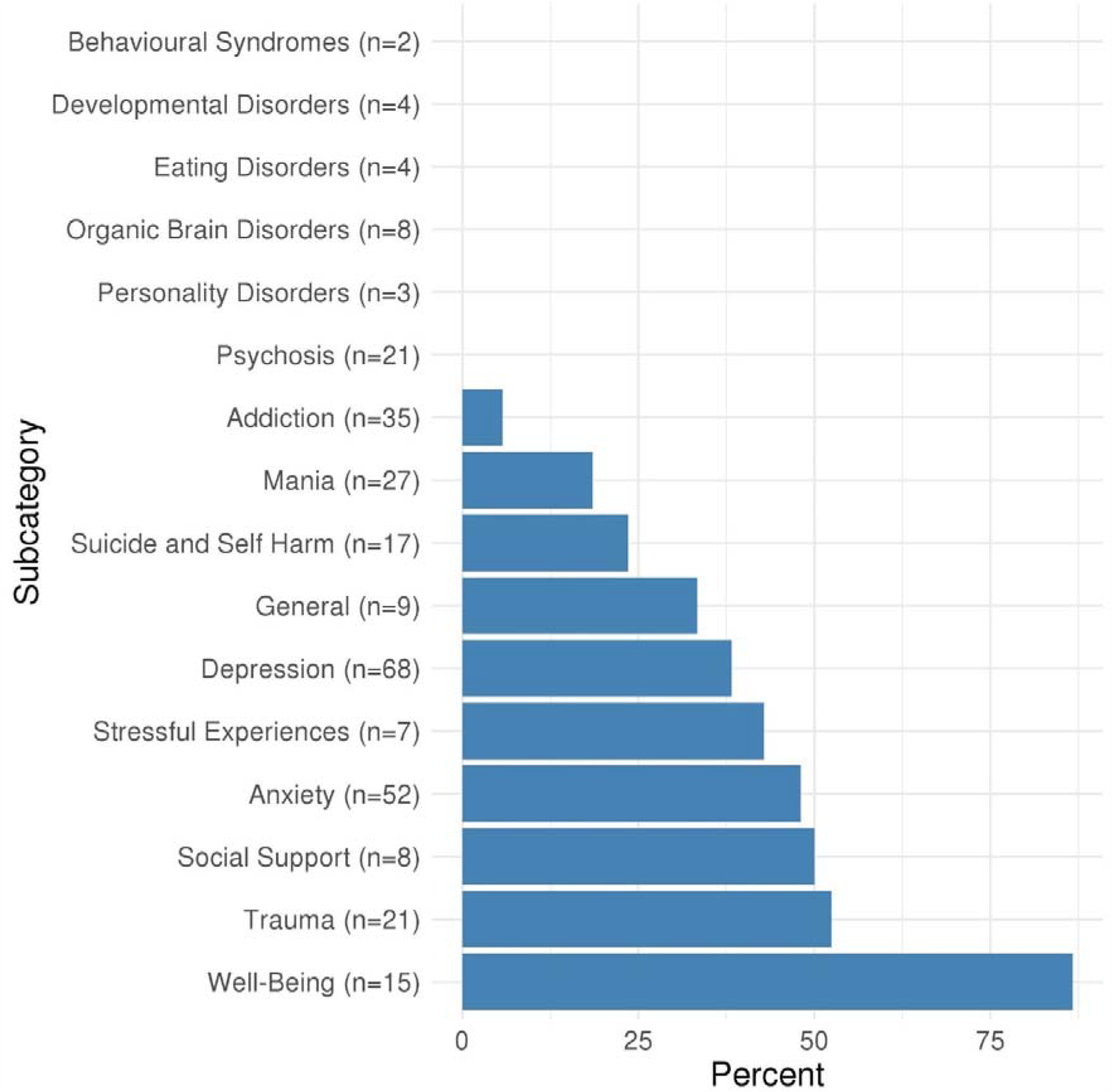
Proportion of potential causal effects of insomnia on outcomes within different mental health subcategories. n is the total number of outcomes in the category. Supplementary Table S3 gives the subcategory for each outcome. Results shown in this figure are also provided in Supplementary Table S5.

Of the physical health category 197 out of 6451 (3%) associations with the insomnia GRS were identified as potential causal effects. Higher proportions of potential causal effects (Figure 4) were seen for the pain (30%) and body composition (19%) subcategories. Examples of adverse potential causal effects on physical health outcomes were long-standing illness, disability or infirmity (OR=1.07, [95% CI: 1.06, 1.07]), back pain in the last month (OR=1.04, [95% CI: 1.04, 1.05]), body fat percentage (mean difference=0.01, [95% CI: 0.01, 0.02]), and gastro-oesophageal reflux (OR=1.05, [95% CI: 1.04, 1.06]).

**Figure 4.**
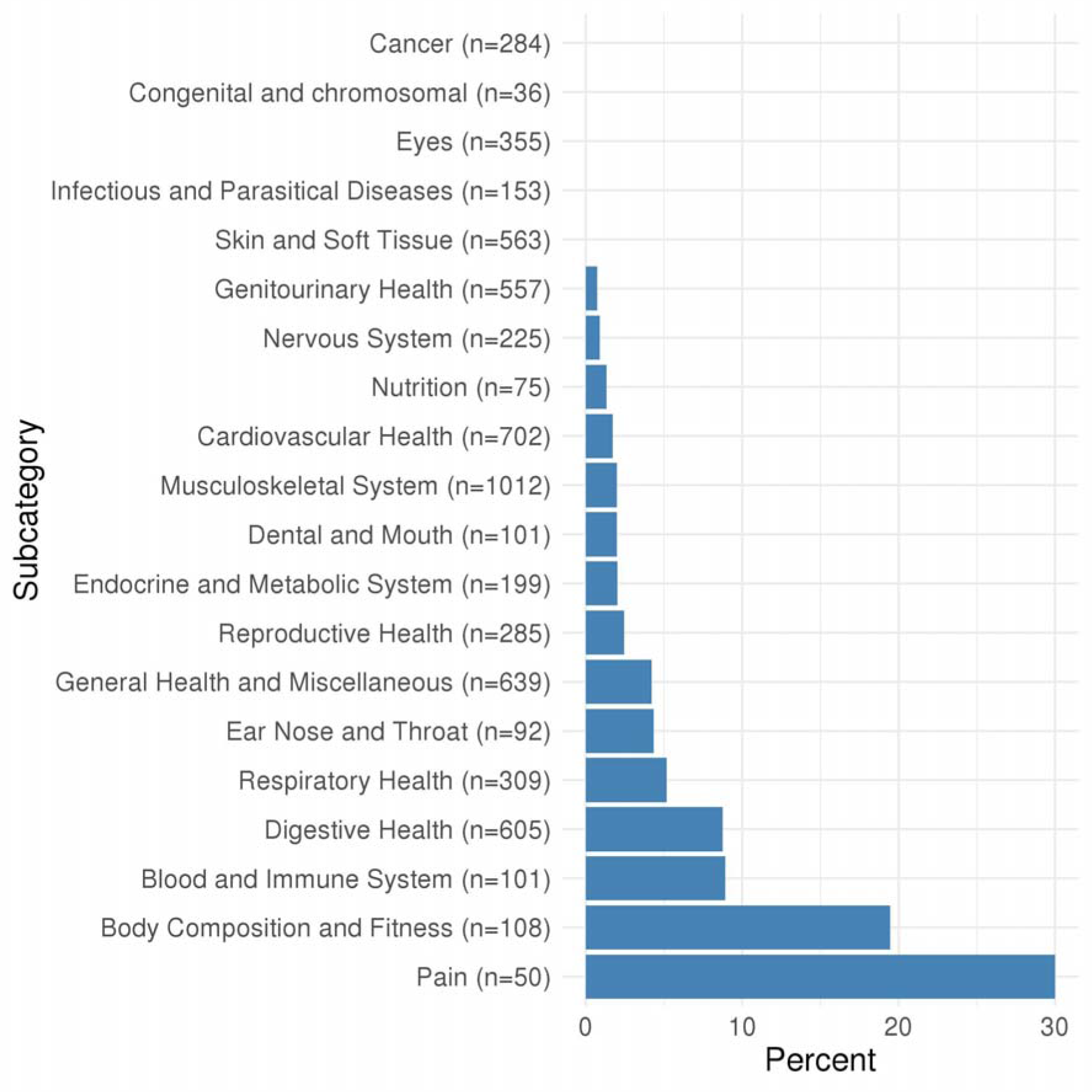
Proportion of potential causal effects of insomnia on outcomes within different physical health subcategories. n is the total number of outcomes in the category. Supplementary Table S3 gives the subcategory for each outcome. Results shown in this figure are also provided in Supplementary Table S5.

For the family and childhood category 17 out of 96 (18%) associations were identified as potential causal effects. This category included some outcomes that could not be plausibly affected by adult insomnia, and might reflect shared family (inherited) predisposition to insomnia and its potential causal effects on fertility and health outcomes across family members, such as maternal smoking around birth (OR=1.03, [95% CI: 1.02, 1.04]), number of full brothers (OR=1.02 [95% CI: 1.02, 1.03]), mother’s age at the time the participant was recruited (mean difference=-0.01, [95% CI: -0.02, 0.01]) and severe depression in a sibling (OR=1.06, [95% CI: 1.04, 1.07]).

Examples of outcomes identified as potential causal effects from the lifestyle/behaviours category (44 out of 854, 5%) are age first had sexual intercourse (mean difference=-0.02, [95% CI: -0.03, -0.02]), time spent watching television (OR=1.04, [95% CI: 1.03, 1.05]) and pack years of smoking (mean difference=0.03, [95% CI: 0.02, 0.03]). Examples of outcomes identified as potential causal effects from the sociodemographic category (38 out of 1053, 4%) are average total household income before tax (OR=0.96, [95% CI: 0.96, 0.97]) and age completed full time education (OR=0.96, [95% CI: 0.95, 0.97]). There were 2 of 2160 (0.1%) outcomes identified as potential causal effects from the brain imaging category. Alternatively, the brain/cognition category had no potential causal effects. Full details of the numbers in each category/subcategory and the numbers and percentages of outcomes in those categories that are potentially influenced by insomnia are provided in Supplementary Tables S4 and S5.

### Sensitivity Analysis

The two GRS used in sensitivity analyses were also associated with an increased risk of insomnia in UK Biobank: OR=1.11 [95%CI: 1.10, 1.12] per one standard deviation increase in the S1 GRS (*p*=1.65×10^−157^, McFadden’s pseudo R^2^=0.01); OR=1.10 [95% CI: 1.10, 1.11] per one standard deviation increase in the S2 GRS (*p*=6.76×10^−145^, McFadden’s pseudo R^2^=0.01). The correlations between each score were strong (R^2^=0.99 [95% CI: 0.99, 0.99] for the S1 GRS with the S2 GRS; R^2^=0.69 [95% CI: 0.69, 0.69] for the S1 GRS with the main GRS; R^2^=0.70 [95% CI: 0.69, 0.70] for the S2 GRS with the main GRS). See Supplementary Figure S2 for associations between each SNP and insomnia for the S1 and S2 GRS.

For GRS S1 and S2, 498 and 490 associations were identified as potential causal effects respectively. There was considerable concordance between the three GRS, with 72% of the 542 associations identified using at least one GRS identified using all three. For associations identified as potential causal effects in any of the three GRS, the association was directionally consistent across all three (Supplementary Figure S3 and Supplementary Table S3).

### Follow-up two-sample MR

Of the 437 potential causal effects identified in the MR-PheWAS, we identified 71 with a relevant GWAS in MR-Base, and hence eligible for follow-up (see Figure 5 and Supplementary Tables S6 – S8). Of these, 36 outcomes showed evidence of an effect of being a self-reported insomnia case versus not (see Supplementary Text for 23andMe definition) in the IVW MR analyses, having 95% confidence intervals which excluded the null (Figures 6 and 7 and Supplementary Tables S9 and S10). These estimates were in the same direction as the MR-PheWAS for 34 of these 36, with 25 (10 continuous and 15 binary) of these having effect estimates in the same direction across all 2-sample MR analyses although with confidence intervals often including the null. These 25 outcomes include a range of categories: mental health outcomes such as anxiety and post-traumatic stress disorder; body composition outcomes such as body fat percentage and waist circumference; musculoskeletal outcomes such as spondylosis, dorsalgia and unspecified arthrosis; the digestive health outcomes diaphragmatic hernia and diseases of the oesophagus, stomach and duodenum; the respiratory outcomes asthma and bronchitis; and outcomes which were not related to others in the set such as C-reactive protein levels, migraine and high-density lipoprotein (HDL) cholesterol. Cochran’s Q showed evidence of between SNP heterogeneity (*p<*0.05) in both the IVW and MR-Egger analyses for nine of these 25 outcomes: asthma, body fat percentage, body mass index, hip circumference, waist circumference, C-reactive protein level, diaphragmatic hernia, migraine and HDL cholesterol, but none showed evidence of horizontal pleiotropy in the MR-egger intercept (see Supplementary Tables S9 and S10 for the Cochran Q and MR-egger results for all outcomes).

**Figure 5.**
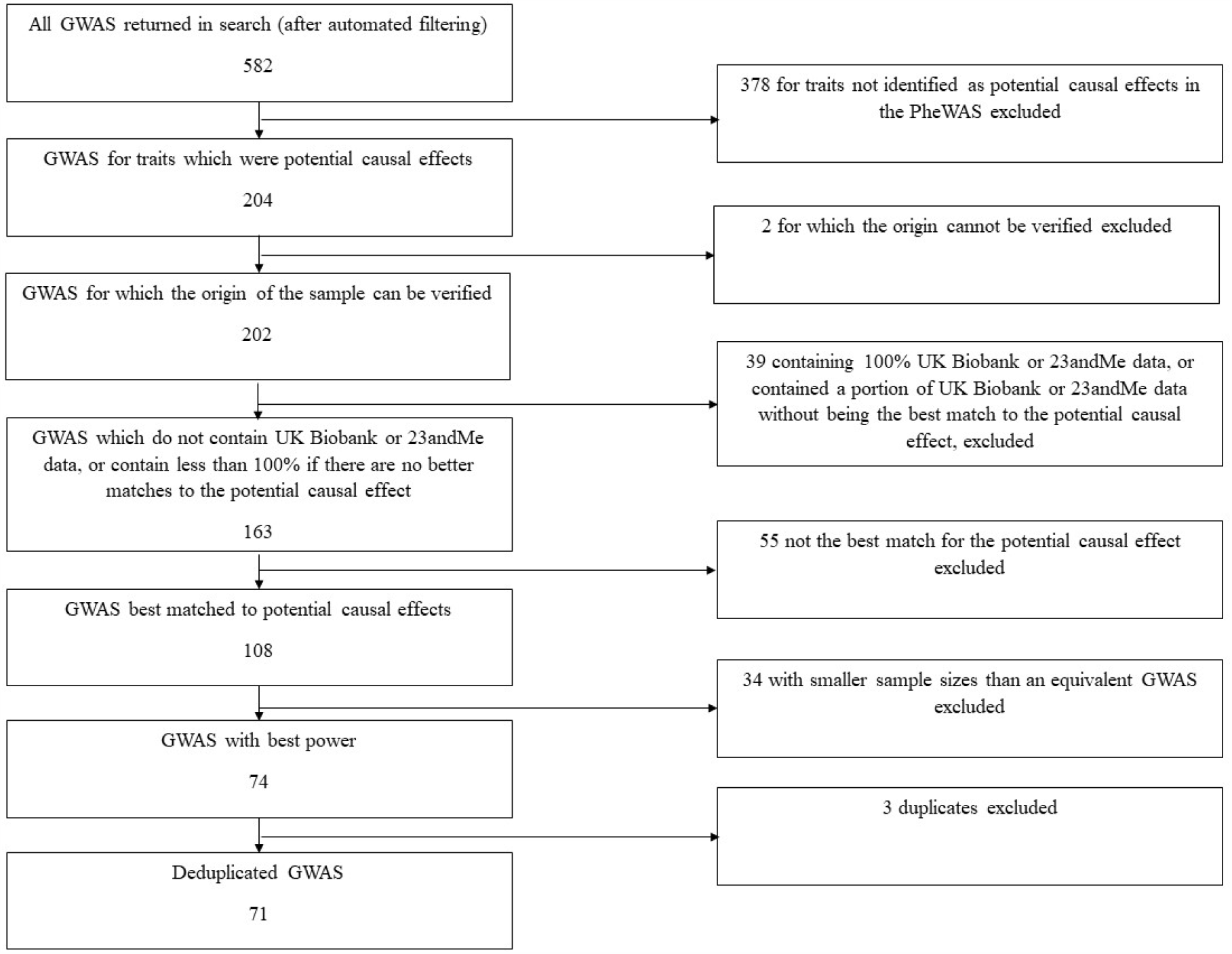
Flow chart of GWAS inclusion for follow-up.

**Figure 6.**
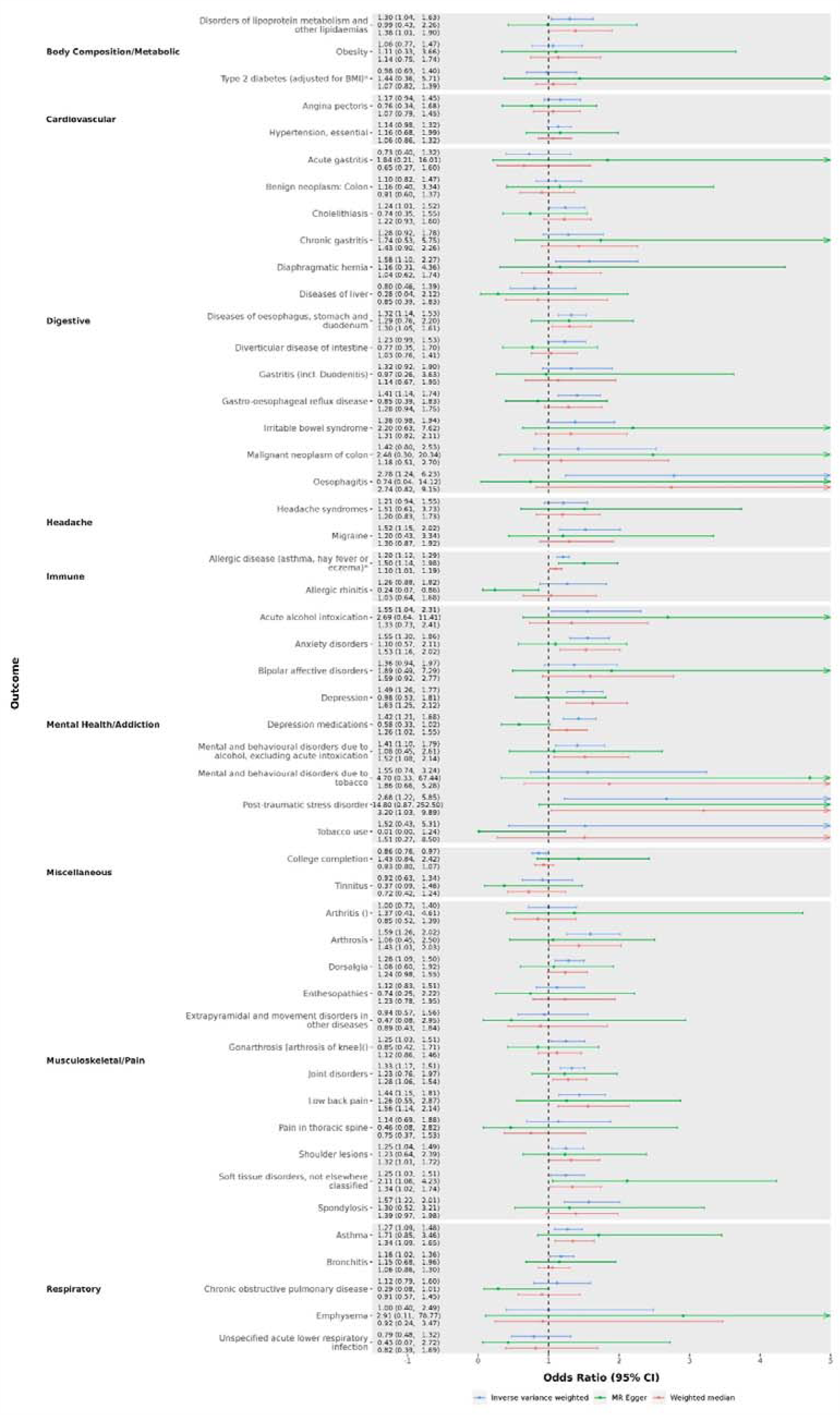
Two sample MR results of the effect (odds ratio), comparing self-reported insomnia cases versus non-cases (see Supplementary Text for 23andMe insomnia definition), for binary outcomes. *GWAS has overlap with UK Biobank or 23andMe (see Supplementary Table S7 for exact percentage).

**Figure 7.**
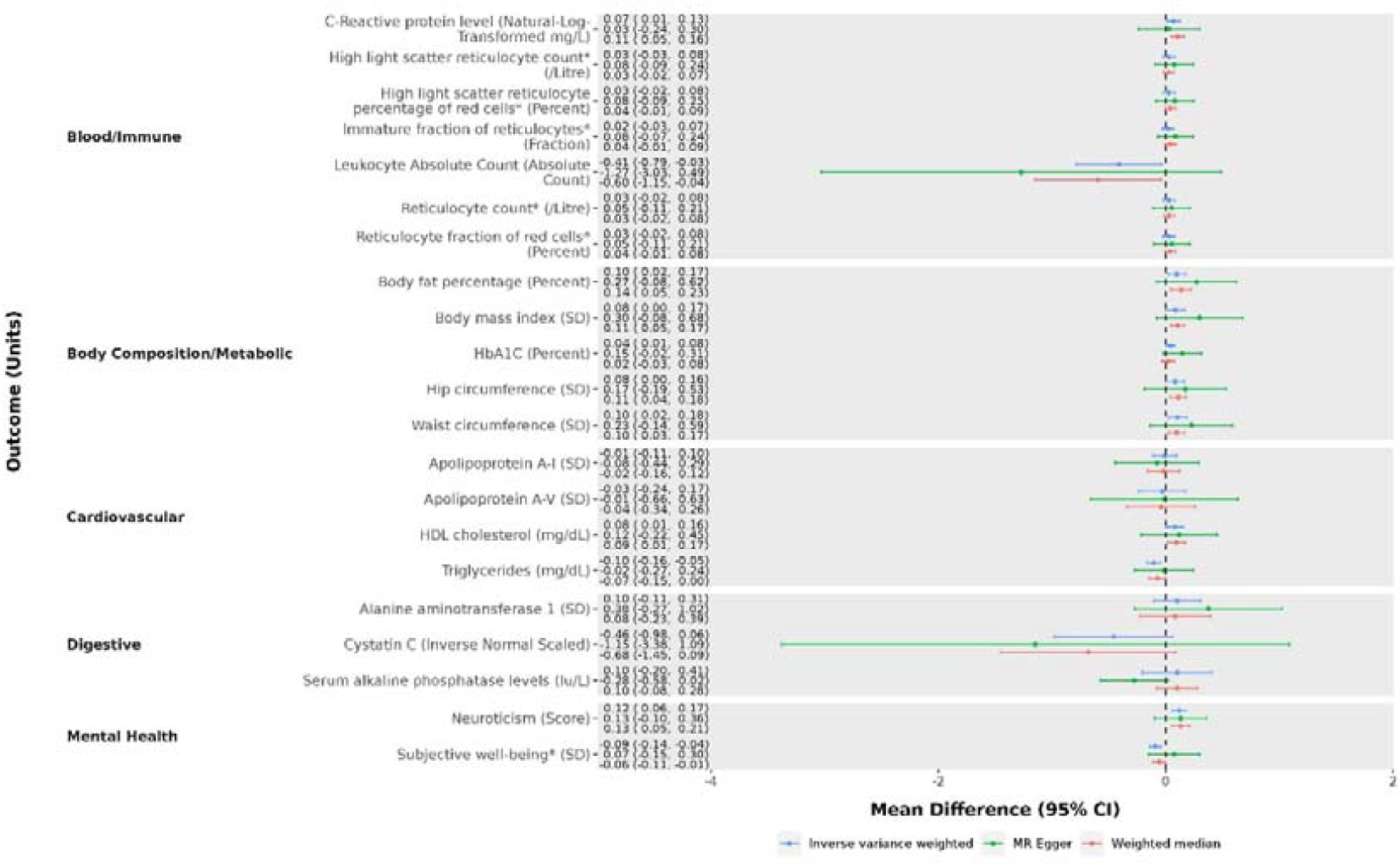
Two sample MR results of the effect (mean difference), comparing self-reported insomnia cases versus non-cases (see Supplementary Text for 23andMe insomnia definition), for continuous outcomes. *GWAS has overlap with UK Biobank or 23andMe (see Supplementary Table S7 for exact percentage).

## Discussion

In this study we conducted an MR-PheWAS of insomnia using a GRS of 129 insomnia-associated SNPs and 11,409 outcome variables, using a subsample of 336,975 unrelated white-British participants from UK Biobank. Of these GRS-outcome associations, 437 met our criteria for being potential causal effects, of which 71 were possible to follow-up using two-sample MR. Follow-up analyses showed consistent evidence of an adverse causal effect of insomnia on 25 outcomes including those related to anxiety disorders, respiratory disorders, musculoskeletal disorders, disorders of the digestive system and body composition measurements. Several of the two-sample MR results were underpowered (sample sizes ranged between 1,000 and 360,838 for outcome GWAS, see Supplementary Table S7) and with larger sample sizes some may be identified as having precisely estimated causal effects of insomnia symptoms. This includes outcomes for which the IVW point estimate was clinically important, such as colon cancer and bipolar disorder. Together with the potential causal effects that we were not able to follow-up, these findings support a role for insomnia in multimorbidity, and the possibility that effective insomnia treatments, such as the cognitive behavioural therapy-Insomnia (31), would prevent a range of other adverse health outcomes. Further work is needed to understand the biological pathways through which the genetic variants may act, and therefore which results are subject to horizontal pleiotropy, as well the mechanisms which might lead to potential causal effects of insomnia on certain health outcomes. Further research is also needed to establish whether the bidirectional causal effects exist between insomnia and the outcomes identified in this work.

Our results indicate that insomnia has numerous and broad effects on health and replicate previous MR studies (in different samples) which indicate insomnia has adverse effects on neuroticism (48), HbA1C levels (increasing) (49), joint and back pain/disorders (27), body composition (24), migraine (50) and alcohol use (51). The finding that lifetime predisposition to insomnia increased C-reactive protein levels (a biomarker for inflammation), aligns with previous experimental research which found a consistent effect of sleep deprivation (52). We also found evidence of novel potential adverse causal effects of insomnia on anxiety disorders, allergic disease (asthma, hay fever or eczema), asthma, bronchitis, soft-tissue disorders, shoulder lesions, diseases of oesophagus/stomach/duodenum, oesophagitis, gastroesophageal reflux disease and diaphragmatic hernia. Evidence was also found for an adverse effect of insomnia on post-traumatic stress disorder, where previous MR studies found no clear evidence (53), and an increasing effect on HDL cholesterol, in the opposite direction to previous MR findings (29).

### Strengths and Limitations

A key strength of our hypothesis-free MR-PheWAS is that it allows for many potential novel causal effects of insomnia to be identified. As multimorbidity is increasingly identified in young and older people, research into how it, and associated polypharmacy, may be prevented, is becoming increasingly important. MR-PheWAS has the potential to identify exposures that could be targets to prevent multimorbidity (8-10). Furthermore, we used two-sample MR to follow-up as many of the potential causal effects as possible and included sensitivity analyses to explore potential bias due to horizontal pleiotropy.

We used a Bonferroni corrected p-value threshold to avoid identifying many potential causal effects that are chance findings, but this may be overly stringent and mean that several outcomes for which insomnia has a true but small causal effect may not have met this threshold due to a lack of statistical power (which differs between outcomes). Also, 366 (84%) potential causal effects could not be followed-up because we were unable to identify a GWAS with summary GWAS data available using MR-Base. Hence, novel potential causal effects still need to be confirmed through further research.

UK biobank has measured a large number of characteristics and has extensive linkage to health records. It also has a large sample size which helps to offset the multiple testing burden of a MR-PheWAS. However, the response rate for UK Biobank was 5.5% and those recruited were on average healthier with lower levels of chronic diseases than the UK population as a whole, which may have resulted in selection bias influencing the MR-PheWAS results (54). Finally, to avoid bias due to population stratification our analyses were restricted to White-Europeans and we cannot assume that our findings would generalise to other ancestral groups.

## Conclusion

Our results suggest that insomnia may have broad effects on health. In particular, we identified novel effects (that replicated in follow-up analyses) on anxiety disorders, allergic disease, respiratory disorders, soft-tissue disorders and digestive disorders, and confirmed previously identified effects on mental health, hyperglycaemia, pain and body composition outcomes. These findings support a role for insomnia in multimorbidity, and the possibility that effective insomnia treatments would prevent a range of other adverse health outcomes. With more and larger GWAS it might be possible to replicate other potential causal effects that we were unable to replicate or obtain precise estimates for.

## Supporting information

Reporting Checklist

## Data Availability

All data is available on request from the UK Biobank or 23andMe. The full GWAS summary statistics for the 23andMe discovery data set will be made available through 23andMe to qualified researchers under an agreement with 23andMe that protects the privacy of the 23andMe participants. Please visit https://research.23andme.com/collaborate/#dataset-access/ for more information and to apply to access the data.

## Footnotes

## Acknowledgements

This research was conducted using the UK Biobank resource under Application Number 16729. This research also used data supplied by 23andMe under a confidentiality agreement. We would like to thank the research participants and employees of 23andMe, Inc. for making this work possible

## Contributor and guarantor information

MJG, DAL and LACM all contributed to the planning of this project. MJG conducted the analysis and wrote the article with supervision and support from DAL and LACM. The corresponding author attests that all listed authors meet authorship criteria and that no others meeting the criteria have been omitted.

## Patient and public involvement reporting

Neither patients or the public were involved at any stage of this study.

## Copyright/license for publication

The Corresponding Author has the right to grant on behalf of all authors and does grant on behalf of all authors, a worldwide licence to the Publishers and its licensees in perpetuity, in all forms, formats and media (whether known now or created in the future), to i) publish, reproduce, distribute, display and store the Contribution, ii) translate the Contribution into other languages, create adaptations, reprints, include within collections and create summaries, extracts and/or, abstracts of the Contribution, iii) create any other derivative work(s) based on the Contribution, iv) to exploit all subsidiary rights in the Contribution, v) the inclusion of electronic links from the Contribution to third party material where-ever it may be located; and, vi) licence any third party to do any or all of the above.

This work carries a Creative Commons Attribution (CC BY 4.0) license.

## Ethical approval

The data collection in UK Biobank was approved by the NHS National Research Ethics Service (ref 11/NW/0382).

## Transparency statement

The lead author affirms that the manuscript is an honest, accurate, and transparent account of the study being reported; that no important aspects of the study have been omitted; and that any discrepancies from the study as originally planned have been explained.

## Supplemental material

This content has been supplied by the author(s). It has not been vetted by BMJ Publishing Group Limited (BMJ) and may not have been peer-reviewed. Any opinions or recommendations discussed are solely those of the author(s) and are not endorsed by BMJ. BMJ disclaims all liability and responsibility arising from any reliance placed on the content. Where the content includes any translated material, BMJ does not warrant the accuracy and reliability of the translations (including but not limited to local regulations, clinical guidelines, terminology, drug names and drug dosages), and is not responsible for any error and/or omissions arising from translation and adaptation or otherwise.

