## Supplementary Text and Figures for "Identifying the potential role of insomnia on multimorbidity: A Mendelian randomization phenome-wide association study in UK Biobank"

Mark Gibson et al.

**Supplementary Text and Figures**

**Supplementary Text**

**UK Biobank genotype data**

UK Biobank genetic data version 3 (2018) was used. The UK BiLEVE Axiom array (n=50,520) and UKB Axiom array (n=438,692), were used for genotyping and covered 812,428 genetic markers. Of these 805,426 markers passed genotype quality controls. Imputation was conducted using the Haplotype Reference Consortium (HRC) reference panel16 (version 1.1), and a reference panel made from the combination of UK10K and the 1000 Genomes project. Imputed data was available for 92,693,895 variants.

**Insomnia Phenotype**

Insomnia cases were identified in UK Biobank (Field 1200) by the participant answering “Usually” to the question “Do you have trouble falling asleep at night or do you wake up in the middle of the night?” as part of the touchscreen questionnaire with participants answering “usually” rather than “never/rarely” or “sometimes” being identified as cases. Participants could also answer with “I do not know” and “Prefer not to answer”. Insomnia cases made up 28% of our final UK Biobank sample.

Insomnia cases were identified in 23andMe by the participant answering yes to one of the following questions:

Have you ever been diagnosed with, or treated for: Insomnia?

Have you ever been diagnosed with, or treated for, any of the following conditions: Insomnia but not Narcolepsy, Sleep apnea or Restless leg syndrome?

Has a doctor ever told you that you have any of these conditions: Insomnia (difficulty getting to sleep or staying asleep)?

Have you ever been diagnosed by a doctor with any of the following neurological conditions: Sleep disturbance?

Do you routinely have trouble getting to sleep at night?

What sleep disorders have you been diagnosed with? Please select all that apply: Insomnia, trouble falling or staying asleep.

Have you ever taken these medications? Prescription sleep aids.

In the last 2 years, have you taken any of these medications? Prescription sleep aids.

Controls were identified by participants answering no to all of the following questions:

Have you ever been diagnosed with, or treated for Insomnia, Narcolepsy, Sleep apnea, Restless leg syndrome?

Have you ever been diagnosed with or treated for any of the following conditions? Post-traumatic stress disorder (PTSD); Autism; Asperger's; Sleep disorder.

Have you ever been diagnosed with or treated for a sleep disorder?

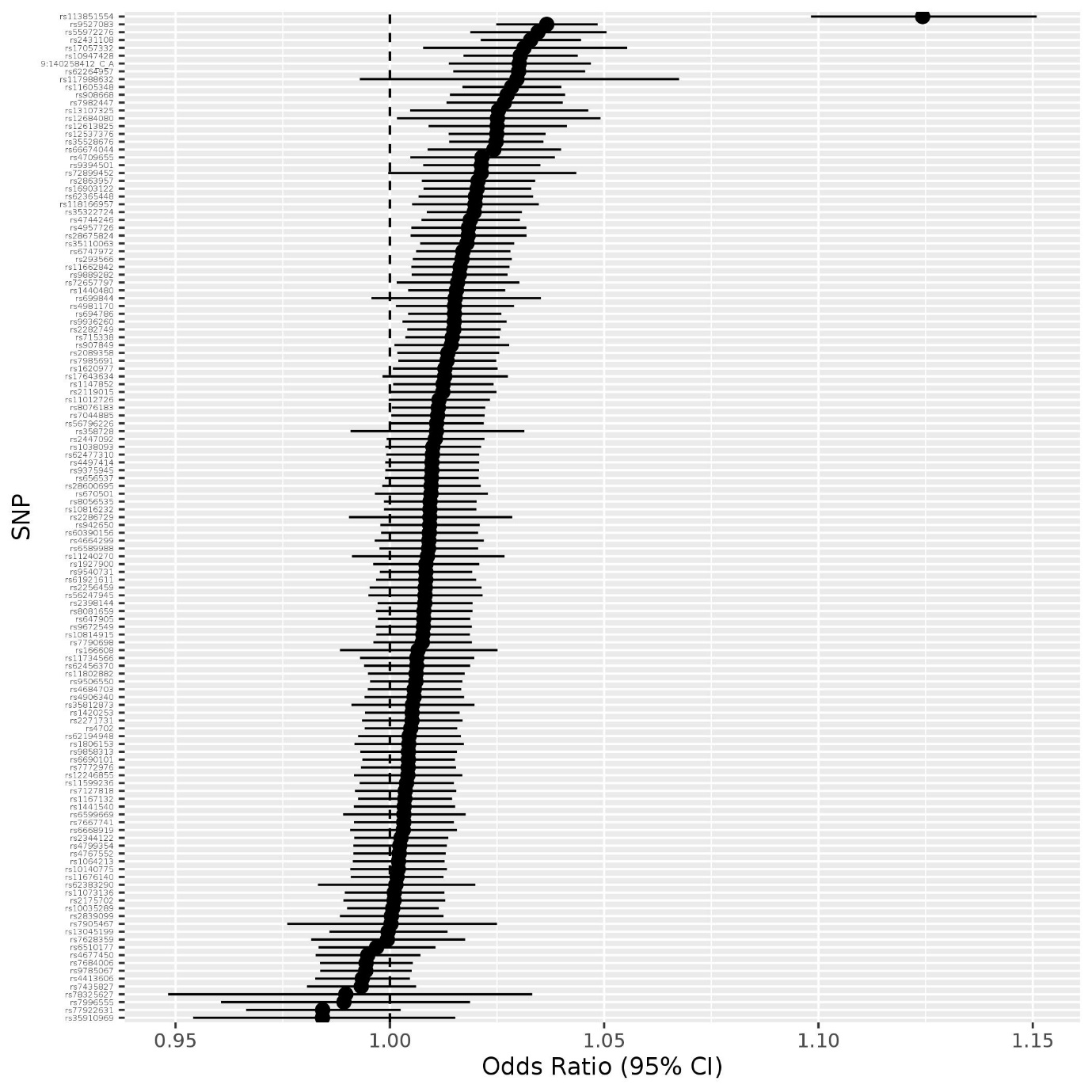

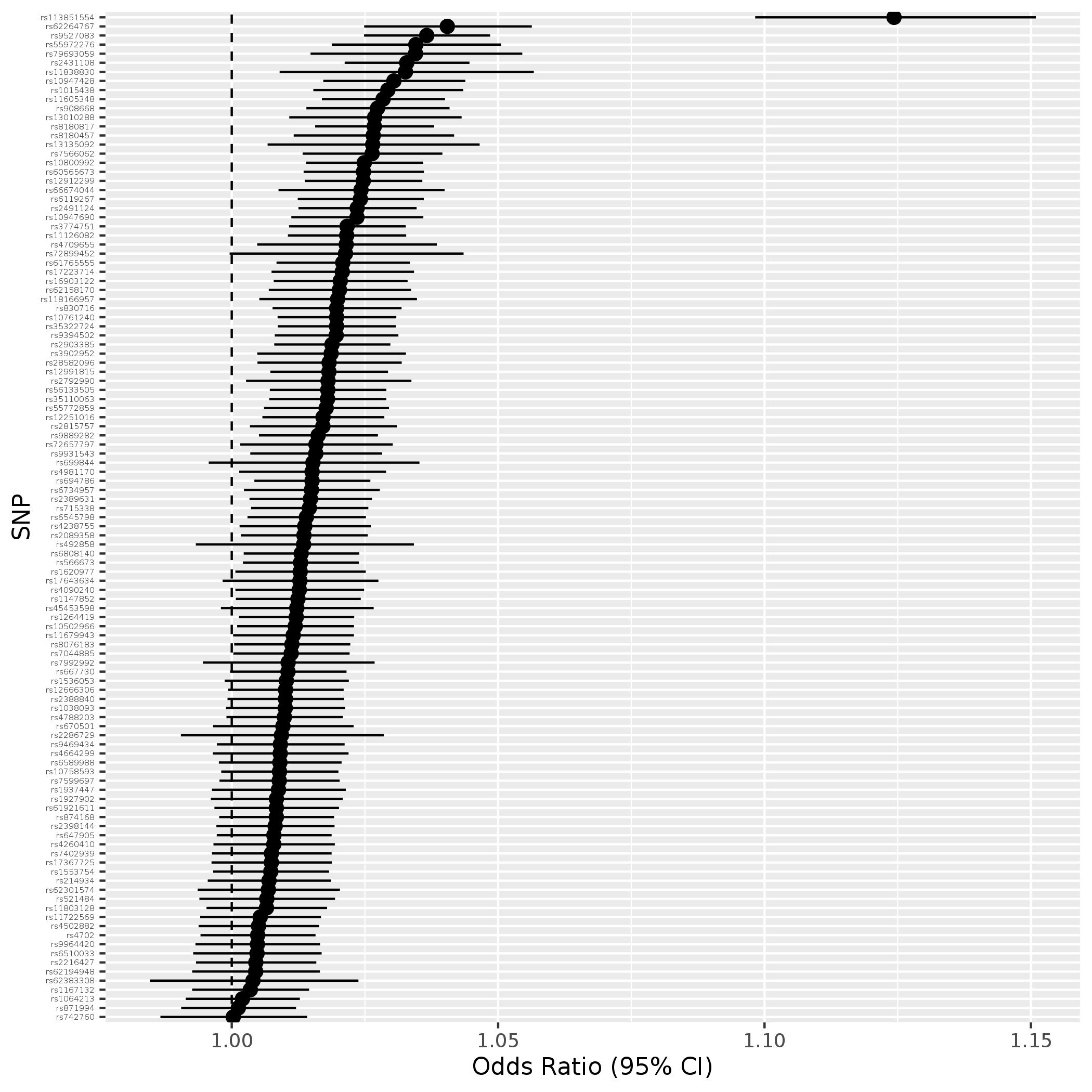

**Supplementary Figure S1**

**Odds ratio and 95% confidence interval for association between each SNP used in the main GRS and insomnia in UK Biobank (Field 1200, with an answer of “usually” coded as an insomnia case).**

**Supplementary Figure S2**

**Odds ratio and 95% confidence interval for association between each SNP used in the S1 and S2 GRS and insomnia in UK Biobank (Field 1200, with an answer of “usually” coded as an insomnia case).**

**S1**: 498 (92%)

**S2**: 490 (90%)

3 (0.6%)

3 (0.6%)

**S1**

10 (2%)

**S2**

2 (0.4%)

93 (17%)

392 (72%)

**Main**

39 (7%)

**Supplementary Figure S3**

**Venn diagram of the number of GRS-outcome associations which passed the Bonferroni-corrected significance threshold for each MR-PheWAS (the percentages are with respect to the total number of associations (542) identified across all MR-pheWAS).**

**Main**: 437 (81%)
